# Predicting and Monitoring Symptoms in Diagnosed Depression Using Mobile Phone Data: An Observational Study

**DOI:** 10.1101/2024.06.15.24308981

**Authors:** Arsi Ikäheimonen, Nguyen Luong, Ilya Baryshnikov, Richard Darst, Roope Heikkilä, Joel Holmen, Annasofia Martikkala, Kirsi Riihimäki, Outi Saleva, Erkki Isometsä, Talayeh Aledavood

**Author notes:** Corresponding Author: Arsi Ikäheimonen, MSc Department of Computer Science Aalto University, FI-02150 Espoo, Finland.

## Abstract

**Background:** Clinical diagnostic assessments and outcome monitoring of patients with depression rely predominantly on interviews by professionals and the use of self-report questionnaires. The ubiquity of smartphones and other personal consumer devices has prompted research into the potential of data collected via these devices to serve as digital behavioral markers for indicating presence and monitoring of outcome of depression.

**Objective:** This paper explores the potential of using behavioral data collected with mobile phones to detect and monitor depression symptoms in patients diagnosed with depression.

**Methods:** In a prospective cohort study, we collected smartphone behavioral data for up to one year. The study consists of observations from 99 subjects, including healthy controls (n=25) and patients diagnosed with various depressive disorders: major depressive disorder (MDD) (n=46), major depressive disorder with comorbid borderline personality disorder (MDD|BPD) (n=16), and bipolar disorder with major depressive episodes (MDE|BD) (n=12). Data were labeled based on depression severity, using the 9-item Patient Health Questionnaire (PHQ-9) scores. We performed statistical analysis and employed supervised machine learning on the data to classify the severity of depression and observe changes in the depression state over time.

**Results:** We identified 32 behavioral markers associated with the changes in depressive state. Our analysis classified depressed subjects with an accuracy of 82% and depression state transitions with an accuracy of 75%.

**Conclusions:** The use of mobile phone digital behavioral markers to supplement clinical evaluations may aid in detecting the presence and relapse of clinical depression and monitoring its outcome, particularly if combined with intermittent use of self-report of symptoms.

## 1. Introduction

In recent years, digital tools and algorithms have become indispensable in healthcare, including mental health. Data-driven technologies have the potential to renew healthcare, providing new avenues for personalized care, remote monitoring, and improved service access. At the same time, mental health disorders, including depression, have remained a significant concern. Depressive disorders are estimated to be the second leading cause of life-years lost to disability worldwide [1]. Alongside markedly impacting individuals’ quality of life, depressive disorders impose a substantial economic burden, including costs to healthcare and societies overall due to disability, reduced employment, and impaired work productivity [2].

Psychiatric evaluations are based on clinical interviews, relying on patients’ self-reflections and recollections, which are susceptible to memory biases and subjective inaccuracies [3]. Further, the absence of definitive physiological biomarkers for mental disorders complicates accurate diagnoses and treatment [4]. Given these challenges, a growing interest has been in data-driven clinical monitoring and decision-making, supplementing subjective evaluations with objective, longitudinal physiological and behavioral data collected via digital devices [5]. This approach, known as digital phenotyping, involves creating a digital representation of a patient’s clinical phenotype using behavioral, social, and physiological markers. The premise of the data-driven approach lies in the inherent value of continuous monitoring, uncovering valuable insights unattainable through intermittent assessments [4].

Recent data-driven studies using devices like smartphones and activity trackers have effectively utilized digital behavioral data to monitor and detect subjects’ depressive moods [6–8]. These studies gather sensor data to identify behavioral patterns associated with depressive disorders such as changes in physical activity, phone usage, and sleep routines. The primary goals include differentiating between depressed patients and healthy controls, classifying mood state transitions, and predicting future mood states. Alongside passively collected data, these studies often use established self-report questionnaires as the reference standard for subjects’ severity of depressive symptoms.

However, some of the studies have employed limited data collection, sample sizes of less than 50 subjects [9–11], a sample of college students [12–15], and data collected over only a few weeks [9,12,16]. Due to these limitations, it may be challenging to generalize results to either a broader population or a free-living setting. Regarding methodologies, earlier research has utilized mobile phone sensors and data categorized as mobile phone usage [9,10,14,15], GPS location data-based features [9–15,17], physical activity data or step counts [11–17], communication patterns [12,14,17], Bluetooth data [13,14], sleep data [13,15], metrics for behavior regularity [15], and physiological measurements [17]. Furthermore, studies have used several metrics for depression as the ground truth, including the 9-item Patient Health Questionnaire (PHQ-9) [18] [9–11,17], a compact version of the 4-item Patient Health Questionnaire (PHQ-4) [19] [15], the Montgomery and Åsberg Depression Rating Scale (MADRS) [20] [16], and the Beck Depression Inventory-II (BDI-II) [21] [13,14]. The analysis methods used in these studies vary, encompassing correlation analysis [9,10,12], machine learning [11,13,14,16,17], and deep learning [15,16].

This paper builds on previous research, exploring the potential of using behavioral data collected with mobile phones to detect and monitor depression symptoms in outpatients diagnosed with depression. Our study aims to identify digital behavioral markers indicative of depressive states and assess the accuracy of this data in detecting depression. Key markers extracted from smartphone sensors, such as the accelerometer, application usage, battery status, communication log, screen activations, and GPS location, comprise metrics like screen-on activation count, total distance traveled, average battery level, phone call count, app usage duration, and maximum acceleration. We analyzed a comprehensive longitudinal dataset, gathered through smartphones, from depressed patients with a diagnosis of either major depressive disorder (MDD), bipolar disorder (MDE|BD), or borderline personality disorder (MDD|BPD) and healthy controls. The focus was on distinguishing subjects self-reporting moderate or more severe depression symptoms and tracking changes in reported depression levels.

## 2. Methods

### Dataset description

We used the data from the Mobile Monitoring of Mood (MoMo-Mood) study, a one-year multimodal digital phenotyping study of individuals undergoing treatment for mental disorders and healthy controls [22]. The MoMo-Mood study recruited 164 participants from four different groups: healthy controls (n=31), patients with major depressive disorder (MDD) (n=85), depressed patients with borderline personality disorder (MDD|BPD) (n=27), and depressed patients with bipolar disorder (MDE|BD) (n=21). Voluntary patients were recruited in Finland from the mood disorder outpatient treatment facilities of the Helsinki University Hospital Mood Disorder Division, Turku University Central Hospital Department of Psychiatry, and City of Espoo Mental Health Services. The patients were diagnosed with structured interviews, namely the Mini-international Neuropsychiatric Interview (MINI) [23] and the Structured Clinical Interview for DSM-IV axis II personality disorders (SCID-II)[24]s, as having ongoing major depressive episodes. Healthy controls were collected by contacting emailing lists of students of the University of Helsinki and Aalto University, users of student health services from these institutions, and recruiting voluntary healthcare personnel from Helsinki University Hospital.

Each group had more females than males: 1) the control group, 24 females, 7 males; 2) the MDD group, 46 females, 31 males; 3) the MDE|BD group, 18 females, 3 males; and 4) the MDD|BPD group, 23 females, 1 male. On average, control group subjects were older than patient group subjects, with average ages as follows: 1) controls 41.8 ± 13.9 years, 2) MDD 39.0 ± 14.2 years, 3) MDE|BD 37.1 ± 10.3 years, and 4) MDD|BPD 28.3 ± 6.0 years. A more detailed description is provided elsewhere [22].

Study participants were recruited on a rolling basis, allowing them to join and leave the research at various intervals. They were requested to stay involved in the study for a maximum of one year; however, they were permitted to withdraw whenever they chose. Consequently, the data gathered on participants vary significantly from several days to one year. The MoMo-Mood study was approved by the Ethics Committee of the Helsinki and Uusimaa Hospital District (HUS) and was granted a research permit by the HUS Department of Psychiatry. As remuneration for their participation, subjects received movie tickets at the end of the initial phase of the study.

Data collection was carried out in two phases. In the initial two weeks, called the *active phase*, participants collected data continuously via personal devices, mobile phones, and actigraphs, and they answered daily mood-related questions. The active phase was followed by the *passive phase,* lasting up to one year. During the passive phase, data collection via mobile phones continued, and participants’ depression was monitored by biweekly PHQ-9 surveys prompted via mobile phone. The PHQ-9 questionnaire comprises nine questions, each scored from 0 to 3, based on the frequency of depressive symptoms over the past two weeks. Thus, the total score ranges from 0 to 27, with high values representing more severe depression. The passive data originate from various mobile phone sensors, including accelerometer, application usage, communication, battery level and screen status logs, and GPS location data. The data were collected through the Niima data collection platform [25]. This work focuses exclusively on the passive phase of the study, employing smartphone data and PHQ-9 survey answers.

### Data Preprocessing

We used Python and Niimpy behavioral data analysis toolbox [26] for data preprocessing. We extracted 93 behavioral features from the raw data. Refer to Table A1 in Multimedia Appendix A for a detailed description of data sources and extracted features. Further, we segmented the data from the accelerometer, application usage, battery status, communication log, and mobile phone screen activations into 6-hour bins (12:00 am to 06:00 am, 6:00 am to 12:00 pm, 12:00 pm to 6:00 pm, and 6:00 pm to 12:00 am). We extracted 308 additional features, resulting in a total of 401 features. The data from different sensors were resampled and averaged using a 14-day sampling. The data were merged with the PHQ-9 questionnaire responses to align data from the preceding biweekly period with the questionnaire responses. Finally, we selected the subjects who had submitted passive data for at least two weeks and answered at least one PHQ-9 questionnaire, yielding 83 subjects.

### Statistical Analysis

#### Distributional testing

To examine whether passively collected mobile phone sensor data show differences between patient groups and control subjects, we employed distributional testing using the non-parametric two-sample Kolmogorov-Smirnov test [27]. The test was chosen due to its capability to detect variations across the entire distribution, including the tails. For the test, we averaged the biweekly sampled data by subject, normalized the data, and omitted the missing values. For robustness against the risk of type I errors (false-positive) due to multiple comparisons, we implemented False Discovery Rate (FDR) correction using the Benjamini-Hochberg procedure [28] at a significance level of α=.05.

#### Correlation analysis

We conducted a correlation analysis to assess the association between passive data features and PHQ-9 scores. We pooled passive data from all subjects, omitted missing values, and applied the Spearman rank correlation coefficient to assess the strength of the relationship. Furthermore, we used FDR correction using the Benjamini-Hochberg procedure at a significance level of α=.05 to account for the multiple testing involved, controlling the expected proportion of false discoveries.

### Predictive Modeling

To achieve the research goal, we deployed supervised machine-learning models for predicting both the presence of depression and state transitions of depressive states. We used a cut-off PHQ-9 depression score of 10 for binary classification analyses. Scores of 10 or higher were considered as *depressed* and scores below 10 as *non-depressed.* We chose a cut-off value of 10 because it signifies clinical depression, typically warranting a treatment plan that may include counseling, follow-up sessions, and possibly pharmacotherapy for the individual. For defining depression state transition, we used the same threshold of 10 and the previous depression state, as presented in Table 1.

**Table 1:** Overview of depression state transition definitions and corresponding labels. This table categorizes the transitions between consequent depression states identified by PHQ-9 scores. Each transition pairs with a specific label, used as a target for the depression state transition modeling.

| Transition description | Label |
| --- | --- |
| Depressed → Depressed | Remains Depressed |
| Depressed → Non-Depressed | Improves |
| Non-Depressed → Non-Depressed | Remains Non-Depressed |
| Non-Depressed → Depressed | Declines |

We built a machine-learning pipeline using Python (version 3.10.8), scikit-learn (version 1.2.0) [29], XGBoost (version 1.7.3) [30], Optuna (version 3.1.0) [31], imbalance-learn (version 0.11.0) [32], and SHAP (version 0.41.0) [33] libraries. Initially, we partitioned our dataset into a 75%/25% train/test split, preventing data leakage by keeping each subject’s data exclusively in either the training or test set. We conducted feature prefiltering by removing features with no or low variance, many missing observations, and a high correlation with other features. We compared filtering and wrapper-based methods and embedded feature selection methods with XGBoost classifiers for feature selection. We used data missingness, variance, and cross-correlation thresholding-based feature selection for the filtering-based method and the Sequential Forward Selection method for the wrapper-based method. Standard preprocessing was applied to selected features, comprising imputation using median values, scaling transformations, and data normalization. To address the class imbalance and improve the robustness of our classification models against overfitting to the majority class, we utilized the Synthetic Minority Oversampling Technique (SMOTE) [34], a method for generating synthetic minority class samples to balance the dataset. We applied SMOTE at two stages of the model’s development. First, during the cross-validation process, the training data folds were balanced using SMOTE. We then applied SMOTE to the entire training dataset in preparation for the final model fitting.

In our study, we focused on the classification task of identifying (1) the presence and (2) the state transitions of depression symptoms using supervised machine learning models. Specifically, three models were examined, namely K-nearest neighbors (KNN), Support Vector Classifier (SVC), and Extreme gradient boosting (XGBoost), all of which are commonly used models in digital phenotyping studies [35,36]. To fine-tune both the model and SMOTE hyperparameters, we employed stratified grouped 5-fold cross-validation, utilizing the Optuna framework [31]. The primary objective in the hyperparameter optimization process was to maximize the F1-score, which balances precision and recall, thereby ensuring a more reliable evaluation of model performance. We also integrated a pruning early stopping technique, which ceases training if there is no improvement in the F1-score (our chosen validation metric). Additionally, the parameters for feature filtering and transformations were optimized during cross-validation.

Finally, we used the test data to evaluate the model performance, assessing the performances with accuracy, precision, recall, negative predictive value (NPV), and F1-scores, as defined in Table 2.

**Table 2:**
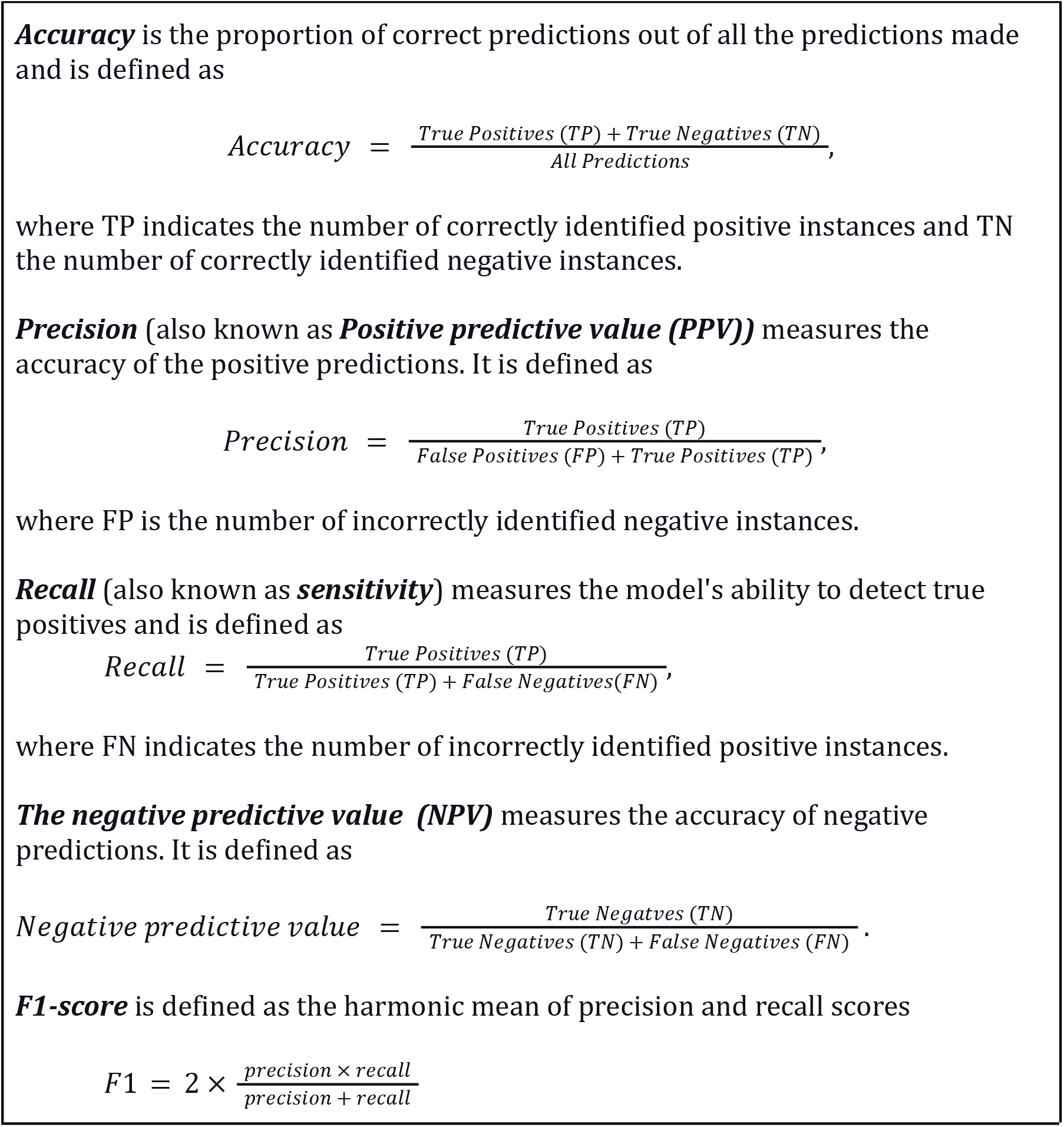
Summary of key performance metrics accuracy, precision, recall, negative predictive value, and F1-score. The table explicates the calculation of these metrics based on the model’s classifications: True Positives (TP) – positive instances correctly predicted, False Positives (FP) – negative instances incorrectly classified as positive, False Negatives (FN) – positive instances incorrectly classified as negative, and True Negatives (TN) – negative instances correctly predicted.

F1-score is a valuable metric because maximizing it ensures that both false positives (identifying a non-depressed subject as depressed) and false negatives (failing to identify a depressed subject) are minimized. High recall reflects low false-negative classification, so we emphasized its importance in model performance evaluation.

### Measuring feature importance

For the final part, focusing on model interpretation, we assessed the importance of features (behavioral markers) for the best-performing XGBoost models to gain insight into the underlying classification mechanisms of the model. We evaluated the importance of each feature for depression presence and the state transition classifications using SHAP (SHapley Additive exPlanations) values [33]. SHAP values measure each feature’s contribution to the model prediction, their relative importance compared with other features, and the significance of feature interactions.

## 3. Results

### Descriptive statistics

The raw data from the passive collection phase contain over 67 million data points, and 819 biweekly PHQ-9 questionnaires gathered data from 99 subjects from 4 subgroups: 25 healthy controls, 46 patients with MDD, 16 patients with MDD|BPD, and 12 patients with MDE|BD. The preprocessing reduced the raw data to 327 200 data points (818 observations with 401 data features) and PHQ-9 scores to 818 observations. The resulting dataset had 83 subjects, comprising 20 healthy controls, 41 MDD patients, 12 patients with MDD|BPD, and 10 patients with MDE|BD.

### PHQ-9 scores

Most of the patients’ PHQ-9 scores during the passive data collection phase remained within the range of 5-19, essentially representing mild to moderate clinical depression, while most control scores remained within the range of 0-4, representing no depression. The group-wise mean scores over the passive phase were as follows: control group 1.2 (±1.8), MDD group 11.9 (±6.7), MDE|BD group 13.7 (±6.5), and MDD|BPD group 13.8 (±6.6). Figure 1 presents these differences, illustrating the distribution of PHQ-9 scores across the various groups.

**Figure 1.**
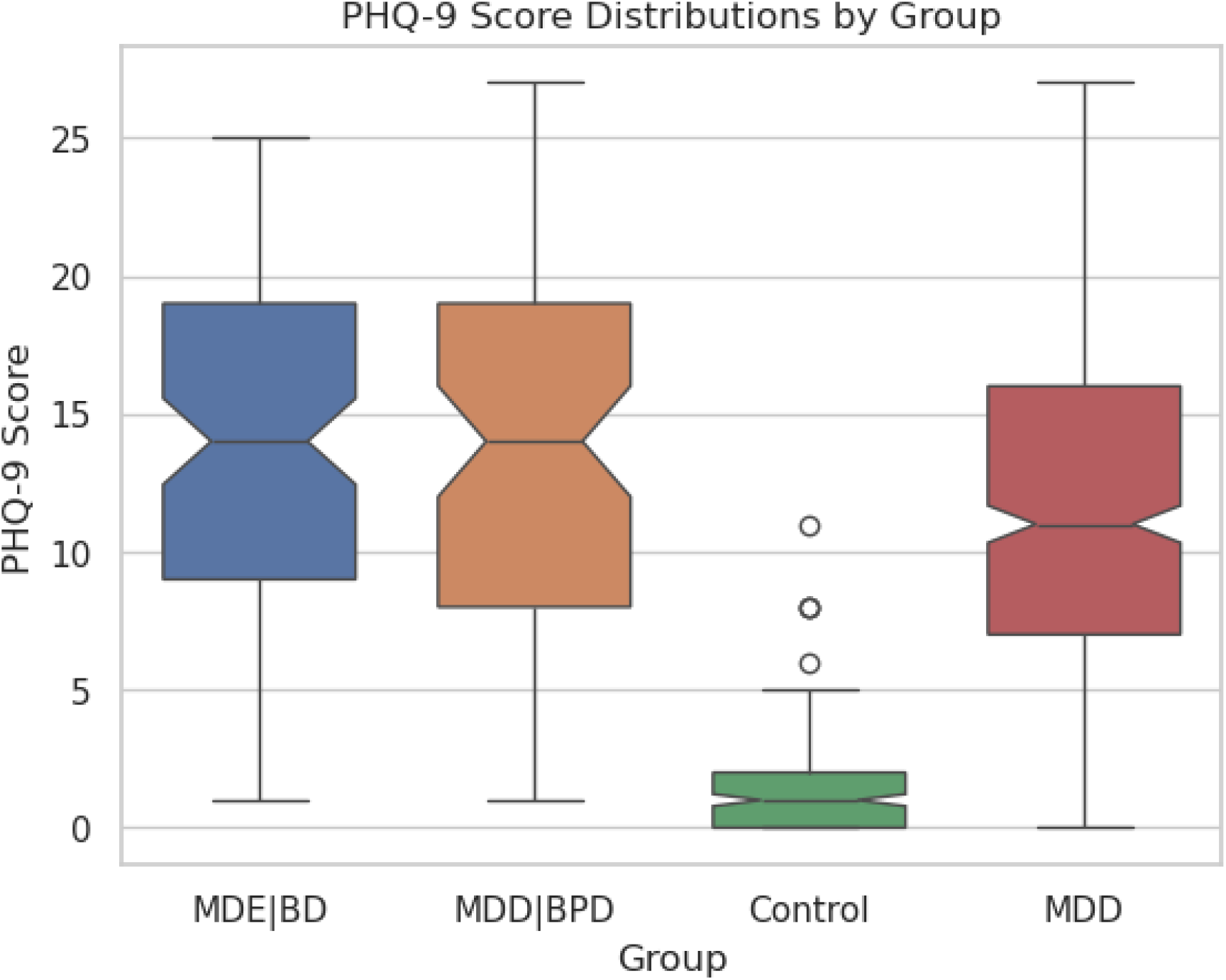
Boxplot representation of PHQ-9 score variability among the control group and three patient groups. Each boxplot displays the interquartile range (IQR), median, and outliers of the PHQ-9 scores. The control group exhibits a smaller median score and less variability, indicating lower and more stable depression symptoms. Conversely, patient groups show higher median scores with a wide IQR, indicating more severe depression and more significant variability in depression severity. The MDD group median is slightly lower than those of the MDE|BD and MDD|BPD groups. It is noteworthy that the patient group scores predominantly represent mild clinical depression.

To assess differences in PHQ-9 scores across various groups, we utilized a Generalized Estimating Equations (GEE) approach [37]. We chose the method due to its effectiveness in dealing with correlated response data and its ability to provide robust standard errors. The analysis revealed statistically significant differences in PHQ-9 scores between the control group and each of the patient groups. The significance of these differences was high, with P<.001 for each comparison.

On average, PHQ-9 scores remain at similar levels within patient groups throughout the study, while all patient groups express a slightly decreasing trend at the beginning of the study. At the group level, MDE|BD and MDD|BPD exhibit more fluctuation in the scores towards the end of the study period, as the number of participants within those groups decreases. Control group scores exhibit a slightly decreasing trend. Figure 2 shows the general group-wise PHQ-9 score trends during the study.

**Figure 2.**
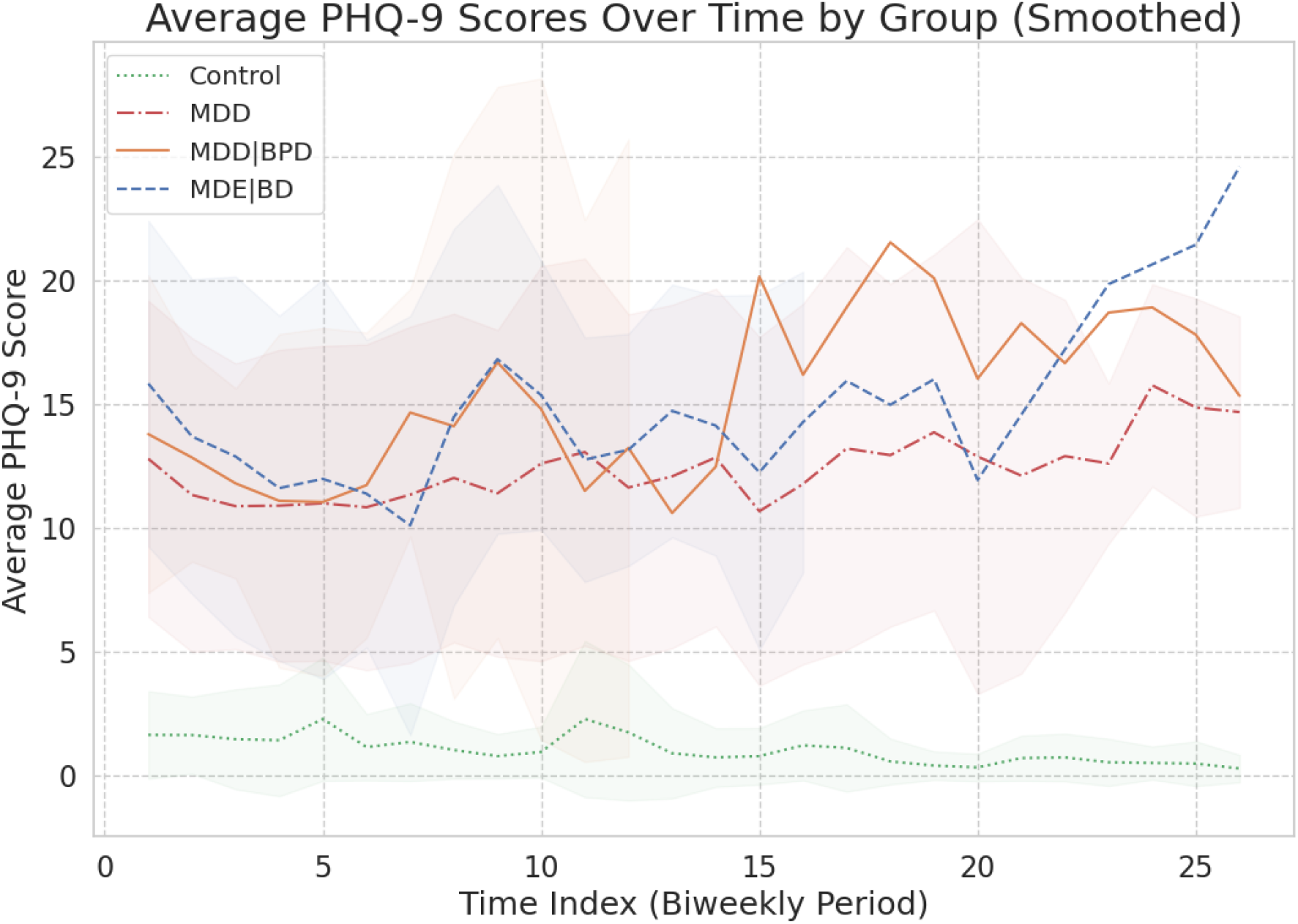
PHQ-9 score trends by group. We averaged the scores within each group, depicted standard deviations with shaded regions, and applied Gaussian smoothing to the line plots for clearer visualization. Patient group average scores have decreasing trends during the first biweekly periods and show increasing trends towards the end of the study. The control group scores remain relatively low, with a minor decreasing trend throughout the study. It is worth noting that the number of participants decreased during the study, increasing the score fluctuations.

We compared the groups by depression severity by categorizing subjects using a cut-off PHQ-9 score of 10 for classifying the subject as depressed or non-depressed. Table 3 shows the cross-tabulated scores for each group.

**Table 3.** Distribution of PHQ-9 scores by severity and group. A PHQ-9 score threshold of 10 was used to categorize individuals into ‘Depressed’ (PHQ-9 score ≥ 10) and ‘Non-Depressed’ (PHQ-9 score < 10) groups. The prevalence of depression severity is shown across different patient groups, with the control group having only one instance of depression.

| Group | Control | MDE\|BD | MDD\|BPD | MDD | All |
| --- | --- | --- | --- | --- | --- |
| **Depression severity** |  |  |  |  |  |
| Depressed | 1 | 65 | 50 | 231 | 347 |
| Non-Depressed | 204 | 36 | 24 | 207 | 471 |
| Total | 205 | 101 | 74 | 438 | 818 |

We categorized the data points into two groups: 347 depressed (42%) and 471 non-depressed (58%) split, resulting in mildly imbalanced classes considering the classification tasks. We assessed biweekly depression state transitions for each group, with detailed descriptions provided in Table 1. Furthermore, Table 4 summarizes these transitions.

**Table 4.** Depression state transitions for the control group and each patient group. The states are classified using a cut-off PHQ-9 score of 10 and comparison with the subject’s previous state. Column ‘Total’ shows the sum of each transition across the groups. Notably, the number of transitions ‘Declines’ and ‘Improves’ is significantly lower than ‘Remains Depressed’ and ‘Remains Non-depressed’.

| Group | Control | MDE\|BD | MDD\|BPD | MDD | Total |
| --- | --- | --- | --- | --- | --- |
| Transition |  |  |  |  |  |
| Declines | 1 | 9 | 5 | 34 | 49 |
| Improves | 1 | 15 | 11 | 43 | 70 |
| Remains Depressed | 0 | 57 | 42 | 222 | 320 |
| Remains Non-depressed | 203 | 20 | 17 | 139 | 379 |
| Total | 205 | 101 | 74 | 438 | 818 |

These results show that in the data the state changes in depression are infrequent compared with the occurrences where the state remains the same. Here, we noticed that transition classes have a significant imbalance, as only 119 (14.5%) state changes counted as transitions and 699 (85.5%) were stationary. This pronounced imbalance could bias classification algorithms towards the majority class, necessitating corrective measures for reliable analysis in subsequent stages.

### Data Completeness

Subject compliance, and thus, data completeness decreased as the study’s passive phase progressed. PHQ-9 survey answer compliance dropped below 70% after six weeks (3 biweekly periods) had passed and thereafter continued to decline steadily. Passive data collection compliance shows a pattern similar to answering the PHQ-9 score questionnaire. Most of the missing data occurs due to the subject dropping out of the study, while some subjects have gaps in data collection. Notably, only a few subjects remained in the study for the entire year. Also, the data collection for some subjects is incomplete due to missing features.

### Statistical Analysis

Two-sample distributional testing using a two-sample Kolmogorov-Smirnov test identified 20 significant features (5%), with *P*-values ranging from .0045 to .0497. However, after applying the FDR correction for multiple comparisons at a significance level α=.05, none of these features were statistically significant; thus, we found no evidence for patient group behavioral data differing from control data. For further details, see Table A2 in Multimedia Appendix A.

Correlation analysis between the behavioral features and PHQ-9 scores using Spearman ranked correlation and FDR correction for multiple comparisons at significance level α=.05 resulted in 32 features (8%) exhibiting statistically significant correlations. The majority (n=18) of the correlations were very weak (absolute value from 0 to 0.19), and the rest (n=14) were weak (absolute value from 0.2 to 0.39). For further details, refer to Table A3 in Multimedia Appendix A.

### Depression Presence Classification

We employed two distinct approaches for classifying the presence of depression. The initial approach treated all biweekly aggregated passive data features (aligned with corresponding biweekly PHQ-9 scores) as independent observations. Utilizing the XGBoost classifier with filter-based feature selection, we achieved the highest accuracy of 66% (95% CI: 56%–70%) and an F1-score of 0.66 (95% CI: 0.5–0.7). The performance comparison of various classifiers and feature selection methods is detailed in Table B1, while Table B2 in Multimedia Appendix B provides a comprehensive summary of the model’s performance.

For the second modeling approach, we included the measured PHQ-9 score from the previous biweekly period as a predictor in the model. Model performance improves notably after adding the predictor. XGBoost classifier with a filtering-based feature selection method achieved the best accuracy of 82% (95% CI: 80%–84%) and a corresponding F1-score of 0.82 (95% CI: 0.80–0.85). This classifier outperformed the other classifiers by a small margin. Table B3 in Multimedia Appendix B summarizes the performance of selected classifiers and feature selection methods, and Table 5 summarizes the XGBoost classifier’s performance.

**Table 5.** Model performance metrics for depression presence classification, using previous PHQ-9 score as a predictor. The model achieved an overall accuracy of 82% across the test data of 208 samples. The table presents the metrics precision, recall, negative predictive value (NPV), and F1-scores for both prediction labels, ‘Non-Depressed’ and ‘Depressed.’ The ‘Support’ column indicates the number of instances for each class in the test dataset. The rows labeled ‘Macro avg’ and ‘Weighted avg’ present the average and support-weighted average values for each metric, respectively. The ‘Non-Depressed’ class (99 samples) achieves precision of 0.80 and recall of 0.83, with an NPV of 0.84 and F1-score of 0.81, reflecting balanced performance. The ‘Depressed’ class (109 samples) has a slightly higher precision of 0.84, a recall of 0.81, an NPV of 0.80, and an F1-score of 0.82, indicating a similar level of predictive accuracy to the ‘Non-Depressed’ class. Both macro and weighted averages across precision, recall, F1-score, and NPV are 0.82, demonstrating consistent performance in detecting both the presence and the absence of depression.

| Metric | Precision | Recall | NPV | F1-score | Support |
| --- | --- | --- | --- | --- | --- |
| Class |  |  |  |  |  |
| Non-Depressed | 0.80 | 0.83 | 0.84 | 0.81 | 99 |
| Depressed | 0.84 | 0.81 | 0.80 | 0.82 | 109 |
| Averages |  |  |  |  |  |
| Macro average | 0.82 | 0.82 | 0.82 | 0.82 | 208 |
| Weighted average | 0.82 | 0.82 | 0.82 | 0.82 | 208 |

### Depression State Transition Classification

For depression state transition classification, we used the XGBoost classifier with feature filtering since it performed best in the depression presence classification. The model was able to classify relatively well the cases where a subject’s state remains the same, while the accuracy is considerably lower for cases where the state changes. Applying SMOTE’s synthetic oversampling technique to alleviate class imbalance significantly increased the recall of the minority classes (depression transitions *’Declines’* and *’Increases’*). Without SMOTE, the model was unable to classify the depression transitions. The best model achieved an accuracy of 75% (95% CI: 72%–76%) and a corresponding F1-score of 0.67 (95% CI: 0.63–0.69). Table 6 summarizes the model validation results. Further, Figure B4 in Multimedia Appendix B displays detailed classification outcomes for the test data.

**Table 6.** Model performance metrics for depression state transition classification. The model’s overall accuracy is 75% for classifying different depression state transitions using the XGBoost classifier with feature filtering. The table presents precision, recall, negative predictive value (NPV), and F1-scores for each transition type, ‘Declines,’ ‘Increases,’ ‘Remains Depressed,’ and ‘Remains Non-Depressed.’ The ‘Support’ column indicates the number of instances for each class in the test dataset, revealing class imbalance with lower counts for ‘Declines’ (17 instances) and ‘Increase’ (23 instances) than for ‘Remains Depressed’ (74 instances) and ‘Remains Non-Depressed’ (94 instances) in a dataset of 208 observations. The macro and weighted (by support) averages are also presented for each metric. For ‘Declines’, the model shows high NPV (0.98) but lower precision (0.34), indicating that while the model reliably identifies cases where the state will not decline, it is less adept at correctly identifying the cases where it declines. The recall is 0.76, leading to an F1-score of 0.47, signifying unbalanced classification performance. ‘Increase’ shows a similar pattern with high NPV (0.96) and moderate recall (0.74) but lower precision (0.46), resulting in an F1-score of 0.57, also indicating unbalanced classification performance. For the ‘Remains Depressed’ and ‘Remains Non-Depressed’ states, the model exhibits higher precision (0.93 and 0.95, respectively) and NPV (0.86 and 0.83, respectively), along with recall rates of 0.72 and 0.77, leading to a more balanced performance with F1-scores of 0.81 and 0.85. The macro average F1 score of 0.67, compared with the overall accuracy of 0.75, reflects the effect of class imbalance on the model’s performance.

| **Metric** | Precision | Recall | NPV | F1-score | Support |
| --- | --- | --- | --- | --- | --- |
| **Transition** |  |  |  |  |  |
| Declines | 0.34 | 0.76 | 0.98 | 0.47 | 17 |
| Increases | 0.46 | 0.74 | 0.96 | 0.57 | 23 |
| Remains Depressed | 0.93 | 0.72 | 0.86 | 0.81 | 74 |
| Remains Non-Depressed | 0.95 | 0.77 | 0.83 | 0.85 | 94 |
| **Averages** |  |  |  |  |  |
| Macro Average | 0.67 | 0.75 | 0.91 | 0.67 | 208 |
| Weighted Average | 0.84 | 0.75 | 0.87 | 0.77 | 208 |

The results show the model’s ability to classify most cases correctly. With an overall accuracy of 75%, the model effectively balances precision across different cases. These findings demonstrate the model’s potential for predicting depression state transitions, leveraging mobile sensed behavioral data and self-reported PHQ-9 scores.

### Feature Importance Analysis using SHAP values

In our analysis of feature importance for classification of presence of depression and depression state transition, we evaluated the relative significance of different features by examining the SHAP values in the best-performing XGBoost models. In summary, our findings highlight the previous PHQ-9 score as the most impactful feature when included in the model. For depression presence classification, additional significant features include mobile phone screen status, application usage, and battery level-related information. In addition to the previous PHQ-9 score for state transition classification, battery level and accelerometer-related features stand out as the most important. Conversely, communication and location-related features had a limited impact on the models.

The importance of the previous PHQ-9 score implies that the depression scores are autocorrelated, thus reflecting future depression level. Mobile phone screen status (e.g. screen on and off event counts) reveals users’ interaction with the device, showing usage frequency and patterns. Similarly, battery level indicates phone usage, reflecting battery drains and charges. Application usage features (especially applications labeled as leisure, sports, and social media) suggest behavioral patterns related to such activities as watching movies or listening to music, exercising, and communicating via social media. Finally, accelerometer-related features reveal physical activity and mobility patterns.

Figures C1–C3 in Multimedia Appendix C present the most important features of these classifications. Specifically, Figure C1 illustrates the important features of depression presence classification without considering the previous biweekly PHQ-9 scores. Conversely, Figure C2 shows the results for the model, including these scores as a predictor. Finally, Figure C3 explores features pertinent to depression state transition classification.

## 4. Discussion

### Principal findings

Our analysis encompassed passively sensed digital behavioral data, which we compared against actively collected PHQ-9 questionnaire data. Employing the Generalized Estimating Equations (GEE) approach, we discovered a statistically significant difference in PHQ-9 score distributions between the control and patient groups. It is important to note that some patients likely experienced recovery post-recruitment for the study, potentially lessening the severity of symptoms reflected in their PHQ-9 scores. Consequently, our data could underrepresent the depression severity spectrum, particularly among patients with more severe depression.

After adjusting for multiple comparisons, distributional testing on behavioral features revealed no significant differences between control and patient groups. This finding suggests that the differences in behavioral data at the group level are minimal. Therefore, our study implies that detecting these subtle differences might require larger sample sizes or alternative statistical methodologies that can leverage hierarchical structures and temporal correlations.

Correlation analysis identified 32 behavioral features with weak or very weak correlations with PHQ-9 scores, predominantly involving mobile phone screen interaction (18 features) and accelerometer data (14 features). Despite most features showing no significant correlation with PHQ-9 scores, their potential value in classification tasks remains, especially considering possible non-linear relationships or interactions with other features.

For the depression prediction tasks, we found that the XGBoost classifier with filtering-based feature selection performed the best in discriminating between depressed and non-depressed subjects, achieving 66% accuracy. The accuracy increased to 82% when we added the PHQ-9 score from the previous biweekly period as a predictor. The difference implies the importance of the temporal structure of the data. Therefore, we propose to include temporal information in future analyses to improve the accuracy. Further, for clinical monitoring applications, information about subjects’ depression history should be available, providing the temporal context necessary to enhance the model’s predictive accuracy.

Furthermore, our results show that the XGBoost classifier, combined with filter-based feature selection and PHQ-9 measurement from the previous biweekly monitoring period as a predictor, can differentiate mood state transitions with a classification accuracy of 75%. While promising, this accuracy level suggests room for further improvement in the model’s performance. Like the depression presence classification, we suggest using more comprehensive methods, personalized models, and temporal information. Additionally, we suspect that the data’s limited sample size and sparsity of transition events hinder the classification performance. Therefore, model development should benefit from a larger sample.

Finally, feature importance analysis revealed insights into the key features of depression prediction models. The most significant predictor for detecting and classifying depression presence was previous biweekly PHQ-9 scores, complemented by features related to accelerometer, application usage, battery level, and screen events. The results emphasize the significance of daily behavioral patterns and time-of-day distinctions (morning, afternoon, evening, and nighttime) in accurately predicting depression. Interestingly, some features were identified with both the correlation and feature importance analyses for classifier models. While the methods and objectives of these analyses differ, the consistency in identifying the same key features across both approaches implies their potential relevance in depression prediction.

### Comparison with previous studies

Our study aligns methodologically with previous research using validated depression assessments and analyzing passively collected smartphone behavioral features. Also, it focuses on statistical inference and machine learning techniques to classify depression among participants and distinguish subjects based on behavioral data. Additionally, the identified important features are consistent with earlier research reporting features related to phone usage [9,10,14,15] and physical activity [11–17]. By contrast, the importance of features related to communication [12,14,17] and location data [9–15,17] were slightly underrepresented in our analysis.

Our classification results are numerically comparable to previous studies using machine learning methods with mobile phone data for depression detection. Using a cohort of college students, Chikersal et al. [14] achieved an 85% accuracy and an F1-score of 0.82 in the post-semester depression detection task. They also achieved an 85% accuracy and an F1-score of 0.80 in detecting a change in the depression state task. Similarly, Wang et al. [15] employed machine learning and deep learning models to detect depression using a subset of mobile phones, also from a cohort of college students, achieving an F1-score of 0.65 using a machine learning model and an F1-score of 0.7 utilizing deep learning.

However, our study differentiates itself by including a diverse cohort of real outpatients, clinically diagnosed with structured interviews, alongside control subjects, thereby offering a broader perspective on depression. Additionally, the data are collected over an extended period in a naturalistic setting, enhancing the reliability of the findings. Unlike other studies that often focus on student populations, it demonstrates the feasibility of digital behavioral monitoring in real outpatients. Furthermore, it excludes certain data features like physiological measurements and social engagement metrics. Lastly, the study does not aim to predict future depressive states, setting it apart from other predictive modeling efforts in the field.

### Limitations

While this research yields insightful outcomes, it is crucial to acknowledge certain limitations. Firstly, dropouts and missing data increase substantially after the first three biweekly periods, limiting our model’s ability to capture patient symptom fluctuations. Secondly, our analysis does not fully account for the hierarchical and autocorrelation structure of the data. We rely on simplified analysis, using aggregated features and pooled subjects, resulting in the loss of available information. Finally, our study does not accommodate external factors that might impact the participants’ behavior patterns and mood states. Given that the data collection partially took place during the COVID-19 era, factors such as social isolation could have played a role in changing the behavior patterns and emotional states of participants.

### Recommendations for future work

This study lays the groundwork for multiple future research endeavors. A direct expansion of our work would be the implementation of personalized models designed to predict the depression state of individuals. These personalized models, which incorporate both group and subject variations and sample-level information, have demonstrated improved accuracy in depression classification tasks [38]. Furthermore, we recommend fully utilizing the temporal structure of the data in classification tasks. Given the inherent variability in symptomatic periods among patients with depression, analyzing temporal patterns and trends from longitudinal data could offer a more accurate representation of their evolving mental states than single-point estimates. We also encourage the exploration of deep learning models in future studies, as these models tend to surpass conventional machine learning methods in predictive accuracy [15,16]. However, due to their complexity and less clear interpretability relative to more traditional methods, we suggest not starting with these models at the outset, instead gradually incorporating them into the analysis. Lastly, to address the challenges posed by the unbalanced dataset in our study, we suggest collecting additional data to enhance the robustness and generalizability of future research findings.

## 5. Conclusions

In summary, this study demonstrates the potential of using mobile phone-sensed behavioral data for monitoring depression symptoms, thereby paving the way for personalized and more effective mental healthcare. The results contribute to an expanding body of evidence supporting the integration of data-driven methods into mental health services. These insights may complement and enhance clinical practices, supplementing conventional diagnostic and monitoring approaches.

## Supporting information

Multimedia Appendix A

Multimedia Appendix B

Multimedia Appendix B

## Data Availability

Due to the highly sensitive and private nature of the data, the data collected in this study cannot be shared with researchers outside of our consortium.

## Acknowledgments

We acknowlege the support and input from Jesper Ekelund in the study design and data collection. The computational resources provided by the Aalto Science-IT project are gratefully acknowledged.

## Abbreviations

BDI-II: Beck Depression Inventory-II
CI: Confidence Interval
FDR: False Discovery Rate
GEE: Generalized Estimating Equations
GPS: Global Positioning System
HUS: Helsinki and Uusimaa Hospital District
KNN: K-Nearest Neighbors
MADRS: Montgomery-Åsberg Depression Rating Scale
MDD: Major Depressive Disorder
MDD|BPD: Major Depressive Disorder with Comorbid Borderline Personality Disorder
MDE|BD: Major Depressive Episodes with Bipolar Disorder
MINI: Mini-International Neuropsychiatric Interview
NPV: Negative Predictive Value
OPTUNA: Optimization Framework
PHQ-4: 4-item Patient Health Questionnaire
PHQ-9: 9-item Patient Health Questionnaire
PPV: Positive Predictive Value
SMOTE: Synthetic Minority Over-sampling Technique
SHAP: SHapley Additive exPlanations
SVC: Support Vector Classifier
XGBoost: Extreme Gradient Boosting

## References

1. World Health Organization. World mental health report: transforming mental health for all. 2022. https://www.who.int/publications/i/item/9789240049338 [accessed Jan 26, 2024].

2. Health TLG. Mental health matters. Lancet Glob Health. 2020;8(11):e1352. doi:10.1016/S2214-109X(20)30432-0

3. Nelson B, McGorry PD, Wichers M, Wigman JT, Hartmann JA. Moving from static to dynamic models of the onset of mental disorder: a review. JAMA Psychiatry. 2017;74(5):528–534.

4. Torous J, Kiang MV, Lorme J, Onnela JP. New tools for new research in psychiatry: a scalable and customizable platform to empower data driven smartphone research. JMIR Ment Health. 2016;3(2):e16.

5. Hsin H, Fromer M, Peterson B, et al. Transforming psychiatry into data-driven medicine with digital measurement tools. NPJ Digit Med. 2018;1(1):1–4.

6. Maatoug R, Oudin A, Adrien V, et al. Digital phenotype of mood disorders: A conceptual and critical review. Front Psychiatry. 2022;13. Accessed February 16, 2023. https://www.frontiersin.org/articles/10.3389/fpsyt.2022.895860

7. Leaning IE, Ikani N, Savage HS, et al. From smartphone data to clinically relevant predictions: A systematic review of digital phenotyping methods in depression. Neurosci Biobehav Rev. 2024;158:105541. doi:10.1016/j.neubiorev.2024.105541

8. Bufano P, Laurino M, Said S, Tognetti A, Menicucci D. Digital Phenotyping for Monitoring Mental Disorders: Systematic Review. J Med Internet Res. 2023;25:e46778. doi:10.2196/46778

9. Saeb S, Zhang M, Karr CJ, et al. Mobile Phone Sensor Correlates of Depressive Symptom Severity in Daily-Life Behavior: An Exploratory Study. J Med Internet Res. 2015;17(7):e4273. doi:10.2196/jmir.4273

10. Saeb S, Lattie EG, Schueller SM, Kording KP, Mohr DC. The relationship between mobile phone location sensor data and depressive symptom severity. PeerJ. 2016;4:e2537.

11. Masud MT, Mamun MA, Thapa K, Lee DH, Griffiths MD, Yang SH. Unobtrusive monitoring of behavior and movement patterns to detect clinical depression severity level via smartphone. J Biomed Inform. 2020;103:103371. doi:10.1016/j.jbi.2019.103371

12. Boukhechba M, Daros AR, Fua K, Chow PI, Teachman BA, Barnes LE. DemonicSalmon: Monitoring mental health and social interactions of college students using smartphones. Smart Health. 2018;9-10:192–203. doi:10.1016/j.smhl.2018.07.005

13. Xu X, Chikersal P, Dutcher JM, et al. Leveraging Collaborative-Filtering for Personalized Behavior Modeling: A Case Study of Depression Detection among College Students. Proc ACM Interact Mob Wearable Ubiquitous Technol. 2021;5(1):41:1–41:27. doi:10.1145/3448107

14. Chikersal P, Doryab A, Tumminia M, et al. Detecting Depression and Predicting its Onset Using Longitudinal Symptoms Captured by Passive Sensing: A Machine Learning Approach With Robust Feature Selection. ACM Trans Comput-Hum Interact. 2021;28(1):3:1–3:41. doi:10.1145/3422821

15. Wang W, Nepal S, Huckins JF, et al. First-Gen Lens: Assessing Mental Health of First-Generation Students across Their First Year at College Using Mobile Sensing. Proc ACM Interact Mob Wearable Ubiquitous Technol. 2022;6(2):95:1–95:32. doi:10.1145/3543194

16. Jakobsen P, Garcia-Ceja E, Riegler M, et al. Applying machine learning in motor activity time series of depressed bipolar and unipolar patients compared to healthy controls. Na KS, ed. PLOS ONE. 2020;15(8):e0231995. doi:10.1371/journal.pone.0231995

17. Mullick T, Radovic A, Shaaban S, Doryab A. Predicting Depression in Adolescents Using Mobile and Wearable Sensors: Multimodal Machine Learning–Based Exploratory Study. JMIR Form Res. 2022;6(6):e35807. doi:10.2196/35807

18. Kroenke K, Spitzer RL, Williams JB. The PHQ-9: validity of a brief depression severity measure. J Gen Intern Med. 2001;16(9):606–613.

19. Kroenke K, Spitzer RL, Williams JBW, Löwe B. An Ultra-Brief Screening Scale for Anxiety and Depression: The PHQ–4. Psychosomatics. 2009;50(6):613–621. doi:10.1016/S0033-3182(09)70864-3

20. Müller MJ, Himmerich H, Kienzle B, Szegedi A. Differentiating moderate and severe depression using the Montgomery–Åsberg depression rating scale (MADRS). J Affect Disord. 2003;77(3):255–260. doi:10.1016/S0165-0327(02)00120-9

21. Beck AT, Steer RA, Brown G. Beck Depression Inventory–II. Published online September 12, 2011. doi:10.1037/t00742-000

22. Baryshnikov I, Aledavood T, Rosenström T, et al. Relationship between daily rated depression symptom severity and the retrospective self-report on PHQ-9: A prospective ecological momentary assessment study on 80 psychiatric outpatients. J Affect Disord. 2023;324:170–174. doi:10.1016/j.jad.2022.12.127

23. Sheehan DV. The Mini-International Neuropsychiatric Interview (M.I.N.I.): The Development and Validation of a Structured Diagnostic Psychiatric Interview for DSM-IV and ICD-10. J Clin Psychiatry.

24. First MB, Benjamin LS, Gibbon M, Spitzer RL, Williams JB. Structured Clinical Interview for DSM-IV Axis II Personality Disorders. American Psychiatric Press; 1997.

25. Aledavood T, Hoyos AMT, Alakörkkö T, et al. Data collection for mental health studies through digital platforms: requirements and design of a prototype. JMIR Res Protoc. 2017;6(6):e110.

26. Ikäheimonen A, Triana AM, Luong N, et al. Niimpy: A toolbox for behavioral data analysis. SoftwareX. 2023;23:101472. doi:10.1016/j.softx.2023.101472

27. Berger VW, Zhou Y. Kolmogorov–Smirnov Test: Overview. In: Wiley StatsRef: Statistics Reference Online. John Wiley & Sons, Ltd; 2014. doi:10.1002/9781118445112.stat06558

28. Benjamini Y, Hochberg Y. Controlling the False Discovery Rate: A Practical and Powerful Approach to Multiple Testing. J R Stat Soc Ser B Methodol. 1995;57(1):289–300. doi:10.1111/j.2517-6161.1995.tb02031.x

29. Pedregosa F, Varoquaux G, Gramfort A, et al. Scikit-learn: Machine Learning in Python. J Mach Learn Res. 2011;12:2825–2830.

30. Chen T, Guestrin C. XGBoost: A Scalable Tree Boosting System. In: Proceedings of the 22nd ACM SIGKDD International Conference on Knowledge Discovery and Data Mining. KDD ‘16. Association for Computing Machinery; 2016:785–794. doi:10.1145/2939672.2939785

31. Akiba T, Sano S, Yanase T, Ohta T, Koyama M. Optuna: A Next-generation Hyperparameter Optimization Framework. In: Proceedings of the 25th ACM SIGKDD International Conference on Knowledge Discovery & Data Mining. KDD ‘19. Association for Computing Machinery; 2019:2623–2631. doi:10.1145/3292500.3330701

32. Lemaître G, Nogueira F, Aridas CK. Imbalanced-learn: A Python Toolbox to Tackle the Curse of Imbalanced Datasets in Machine Learning. J Mach Learn Res. 2017;18(17):1–5. Accessed December 5, 2023. http://jmlr.org/papers/v18/16-365.html

33. Lundberg SM, Lee SI. A unified approach to interpreting model predictions. Adv Neural Inf Process Syst. 2017;30.

34. Chawla NV, Bowyer KW, Hall LO, Kegelmeyer WP. SMOTE: Synthetic Minority Over-sampling Technique. J Artif Intell Res. 2002;16:321–357. doi:10.1613/jair.953

35. Benoit J, Onyeaka H, Keshavan M, Torous J. Systematic Review of Digital Phenotyping and Machine Learning in Psychosis Spectrum Illnesses. Harv Rev Psychiatry. 2020;28(5):296. doi:10.1097/HRP.0000000000000268

36. Jacobson NC, Summers B, Wilhelm S. Digital Biomarkers of Social Anxiety Severity: Digital Phenotyping Using Passive Smartphone Sensors. J Med Internet Res. 2020;22(5):e16875. doi:10.2196/16875

37. Hardin JW, Hilbe JM. Generalized Estimating Equations: GEE. 2. ed. Chapman & Hall/CRC; 2013.

38. Palmius N, Saunders KEA, Carr O, Geddes JR, Goodwin GM, Vos MD. Group-Personalized Regression Models for Predicting Mental Health Scores From Objective Mobile Phone Data Streams: Observational Study. J Med Internet Res. 2018;20(10):e10194. doi:10.2196/10194

