## Supplementary material for "Predicting and Monitoring Symptoms in Diagnosed Depression Using Mobile Phone Data: An Observational Study": Multimedia Appendix A

**Figure A1.** Summary of data sources and extracted features for each sensor. This table summarizes the data sources, specifically the sensors used, and the corresponding features extracted from each sensor. For a detailed reference, see Niimpy documentation (“Niimpy: Behavioral data analysis”, 2023).

| **Sensor** | **Extracted Features** |
| --- | --- |
| Accelerometer | Acceleration magnitude, magnitude max, min, mean and standard deviation, measurement count |
| Application | Application class, count, duration |
| Battery | Battery level mean, median and standard deviation, battery shutdown time, battery discharge |
| Communication | Total call duration, call duration mean, median and standard deviation, call count, outgoing-incoming call ratio, SMS count |
| Location | *Distance-based features:* total distance, variance, log variance, average speed, speed variance, max speed, location bin count.  *Significant place-related features:* static point count, moving point count, static bin count, max distance from home, number of significant places, number of rarely visited places, number of transitions between significant places, bin count in top1, top2, top3, top4, and top5 cluster, normalized entropy |
| Screen | Screen off timestamp, screen event count, screen event duration, screen event duration max, mean, min, and standard deviation, screen first unlock timestamp |
| Survey | PHQ-9 score |

**Figure A2.** Kolmogorov-Smirnov test results for behavioral data distribution differences. This table presents the Kolmogorov-Smirnov(KS) -test results, assessing the behavioral data distribution differences between the control and patient groups, MDE|BD, MDD|BPD, and MDD. Features are included in this table if they demonstrate any statistical difference between at least one of the three groups and the control group. Importantly, although differences are observed (20 features out of 401), none of the features reached statistical significance (p < .05) after controlling the false discovery rate with the Benjamini-Hochberg procedure. The presented features were extracted from the smartphone accelerometer, application usage, battery level, and screen events data.

| **Group** | **MDE\|BD** |  | **MDD\|BPD** |  | **MDD** |  |
| --- | --- | --- | --- | --- | --- | --- |
|  | **P-val** | **Corrected p-val** | **P-val** | **Corrected p-val** | **P-val** | **Corrected p-val** |
| **Feature** |  |  |  |  |  |  |
| Morning acceleration standard deviation | .01 | .36 | .03 | .94 | .034 | >.999 |
| Leisure applications usage count | < .001 | .69 | .38 | >.999 | .31 | .87 |
| Sports applications usage count | .72 | >.999 | .004 | .37 | .14 | .87 |
| Afternoon leisure appilcations usage count | .04 | .8 | .47 | >.999 | .18 | .87 |
| Morning leisure appilcations usage count | .03 | .61 | .08 | >.999 | .06 | >.999 |
| Nighttime security applications usage count | .48 | >.999 | .02 | .77 | .6 | >.999 |
| Afternoon mean battery level | .17 | .92 | .53 | >.999 | .02 | >.999 |
| Morning mean battery level | .02 | .43 | .31 | >.999 | .05 | >.999 |
| Afternoon acceleration count | .17 | .92 | .02 | .77 | .17 | .87 |
| Nighttime acceleration count | .01 | .38 | .003 | .37 | .06 | >.999 |
| Evening maximum acceleration | .57 | >.999 | .05 | >.999 | .13 | .87 |
| Morning maximum accelration | .02 | .39 | .03 | .94 | .02 | >.999 |
| Afternoon minimum acceleration | .01 | .83 | .18 | >.999 | .59 | >.999 |
| Evening minimum acceleration | .01 | .36 | .21 | >.999 | .62 | >.999 |
| Nighttime minimum acceleration | .09 | .87 | .15 | >.999 | .01 | >.999 |
| Afternoon mean acceleration | .09 | .87 | .01 | .59 | .03 | >.999 |
| Nighttime mean accelaration | .14 | .9 | .04 | >.999 | .009 | >.999 |
| Morning screen use count | .04 | .74 | .07 | >.999 | .11 | .87 |
| Leisure applications usage duration | .01 | .36 | .79 | >.999 | .30 | .87 |
| Sports applications usage duration | .72 | >.999 | .009 | .59 | .17 | .87 |

**Figure A3.** Spearman rank correlation test results between behavioral data features and PHQ-9 Scores. This table presents the Spearman rank correlation test results between the behavioral data features and the biweekly assessed PH9-scores. Only the features exhibiting statistically significant correlation (32 out of 401) are shown in the table. Column ‘correlation’ shows the test correlation coefficient, ‘P-value’ corresponding P-value, and ‘corrected P-value’ the P-values after applying the Benjamini-Hochberg procedure to control type-I errors due to multiple comparisons at the significance level of α=.05. These presented features are extracted from smartphone accelerometer and screen activations. The correlation coefficients range from -0.38 to 0.17, thus ranging from low to moderate correlations.

| **Feature** | **Spearman correlation** | ***P*-value** | **Corrected *P*-value** |
| --- | --- | --- | --- |
| Nightltime screen usage count | 0.17 | <.001 | <.001 |
| Nighttime acceleration standard deviation | 0.14 | <.001 | <.001 |
| Morning minimum acceleration | 0.13 | <.001 | <.001 |
| Afternoon screen usage count | 0.11 | <.001 | <.001 |
| Screen-on total duration | 0.11 | <.001 | .01 |
| Nighttime screen-off count | 0.1 | .01 | .01 |
| Nighttime screen usage duration standard deviation | 0.09 | .01 | .03 |
| Morning screen usage count | -0.08 | .02 | .04 |
| Evening screen usage duration standard deviation | -0.11 | <.001 | .01 |
| Morning maximum screen usage duration | -0.11 | <.001 | <.001 |
| Evening minimum acceleration | -0.12 | <.001 | .01 |
| Evening acceleration count | -0.12 | <.001 | .01 |
| Morning screen-off count | -0.12 | <.001 | <.001 |
| Morning screen usage duration standard deviiation | -0.13 | <.001 | <.001 |
| Afternoon maximum acceleration | -0.14 | <.001 | <.001 |
| Morning acceleration count | -0.16 | <.001 | <.001 |
| Acceleration count | -0.19 | <.001 | <.001 |
| Afternoon acceleration | -0.2 | <.001 | <.001 |
| Afternoon minimum screen-off duration | -0.22 | <.001 | <.001 |
| Morning median screen-off duration | -0.22 | <.001 | <.001 |
| Evening acceleration | -0.22 | <.001 | <.001 |
| Afternoon maximum screen-off duration | -0.22 | <.001 | <.001 |
| Nighttime acceleration count | -0.23 | <.001 | <.001 |
| Morning maximum screen-off duration | -0.23 | <.001 | <.001 |
| Afternoon median screen-off duration | -0.24 | <.001 | <.001 |
| Morning screen-off duration standard deviation | -0.27 | <.001 | <.001 |
| Morning maximum acceleration | -0.27 | <.001 | <.001 |
| Total screen-off duration | -0.29 | <.001 | <.001 |
| Morning acceleration | -0.3 | <.001 | <.001 |
| Nighttime acceleration | -0.33 | <.001 | <.001 |
| Evening maximum screen-off duration | -0.38 | <.001 | <.001 |
| Nighttime minimum acceleration | -0.38 | <.001 | <.001 |
