## Supplementary material for "Predicting and Monitoring Symptoms in Diagnosed Depression Using Mobile Phone Data: An Observational Study": Multimedia Appendix B

**Table B1.** Performance Comparison of Depression Presence Classification Models**.** This table presents a performance comparison of depression presence classification models. The table presents the model accuracy and F1-scores for three classifiers, XGBoost, KNN, and SVC, across different feature selection methods: filter-based, wrapper-based, and without feature selection. The highest performance metrics for each classifier are highlighted in bold. Specifically, the XGBoost classifier with filter-based feature selection achieved the best overall accuracy at 66% and an F1-score of 0.66. The results show that feature selection with a feature filtering method benefits all classifier models.

| **Feature Selection Method** | Filter |  | Wrapper |  | Without feature selection |  |
| --- | --- | --- | --- | --- | --- | --- |
| **Metric** | Accuracy | F1-score | Accuracy | F1-score | Accuracy | F1-score |
| **Classifier** |  |  |  |  |  |  |
| **XGBoost** | **0.66** | **0.66** | 0.62 | 0.60 | 0.61 | 0.60 |
| **KNN** | **0.57** | **0.57** | 0.49 | 0.49 | 0.50 | 0.48 |
| **SVC** | **0.62** | **0.62** | 0.48 | 0.46 | 0.54 | 0.53 |

**Table B2.** Performance metrics for depression presence classification: Non-Depressed vs. Depressed. This table presents the performance metrics for a depression presence classification model distinguishing between two classes: Non-Depressed and Depressed. The model demonstrates a moderate accuracy of 66% across 208 samples. The 'Support' column indicates the number of instances for each class in the test dataset. For Non-Depressed, the precision is 0.63, indicating that 63% of predictions for Non-Depressed were correct, while the recall of 0.73 shows that 73% of actual Non-Depressed cases were correctly identified. The NPV value of 71% means that when the model predicts someone is not depressed, there is a 71% chance they are genuinely not depressed. The F1-score for Non-Depressed is 0.67, affected by the lower precision score. For Depressed, the model achieves a precision of 0.71, a recall of 0.61, an F1-score of 0.65, and an NPV of 0.63. The table also presents macro and weighted averages for precision, recall, F1-score, and NPV, ranging between 0.66 and 0.67, indicating a balanced performance between the two classes. Overall, the model exhibits a marginally higher accuracy in correctly identifying Non-Depressed cases than Depressed cases. The precision and recall metrics imply that the model is more conservative in predicting instances of depression. However, the moderate NPV values for both classes indicate room for improvement, especially in minimizing false negatives, which would be critical in a clinical setting.

| **Metric** | Precision | Recall | NPV | F1-score | Support |
| --- | --- | --- | --- | --- | --- |
| **Depression Severity** |  |  |  |  |  |
| Non-Depressed | 0.63 | 0.73 | 0.71 | 0.67 | 99 |
| Depressed | 0.71 | 0.61 | 0.63 | 0.65 | 109 |
| **Average** |  |  |  |  |  |
| Macro Average | 0.67 | 0.67 | 0.67 | 0.66 | 208 |
| Weighted Average | 0.67 | 0.66 | 0.67 | 0.66 | 208 |

**Table B3.** Comparative performance of depression classification models with biweekly PHQ-9 score as a predictor. This table presents a performance comparison of depression presence classification models, including the previous biweekly PHQ-9 Score as a predictor. The table presents the model accuracy and F1-scores for three classifiers, XGBoost, KNN, and SVC, across different feature selection methods: filter-based, wrapper-based, and without feature selection. The highest performance metrics for each classifier are highlighted in bold. Specifically, the XGBoost classifier with filter-based feature selection achieved the best overall accuracy at 82% and an F1-score of 0.82. The results show that feature selection benefits all models.

| **Feature selection method** | Filter |  | Wrapper |  | Without feature selection |  |
| --- | --- | --- | --- | --- | --- | --- |
| **Metric** | Accuracy | F1-score | Accuracy | F1-score | Accuracy | F1-score |
| **Classifier** |  |  |  |  |  |  |
| XGBoost | **0.82** | **0.82** | 0.79 | 0.79 | 0.77 | 0.77 |
| KNN | 0.74 | 0.73 | **0.76** | **0.76** | 0.63 | 0.57 |
| SVC | 0.73 | 0.71 | **0.80** | **0.80** | 0.71 | 0.70 |

**Figure B4**. Confusion matrix for depression state transition classification. This confusion matrix represents the validation results of a model used for classifying depression state transitions. The matrix displays the validation results for the model classifying depression state transitions, contrasting true labels (shown on the y-axis) with the model's predictions (on the x-axis). The matrix cells display the count of predictions for each state, with diagonal cells representing correct predictions. While the matrix illustrates the model's ability to correctly identify most of the 'remains depressed' and 'remains non-depressed' cases, it also reveals a tendency to misclassify them as other transitions. Both 'declines' and 'increases' states show some misclassifications, reflecting challenges in the model's performance with less frequent states.

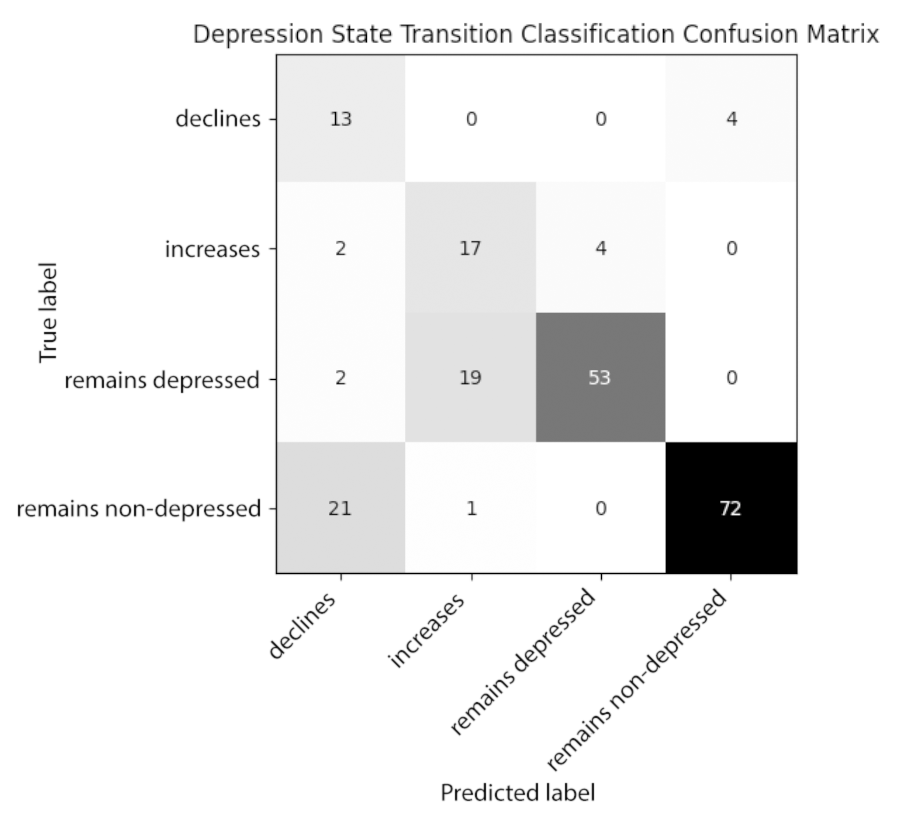
