## Supplementary material for "Predicting and Monitoring Symptoms in Diagnosed Depression Using Mobile Phone Data: An Observational Study": Multimedia Appendix B

### **Multimedia Appendix C**

**Figure C1.** Key features in depression presence classification model based on SHAP values**.** This bar chart illustrates the most important features of a depression presence classification model, as evaluated using SHAP values. The chart orders the features by the magnitude of their average impact on the model's predictions, with a higher mean SHAP value indicating a higher influence on the model's output. 'Screen-off duration standard deviation' (indicating the fluctuation in smartphone screen-off durations) is the most important feature, followed by 'Longest evening screen-off duration' (indicating the longest period of smartphone screen turned off during the evening), 'Leisure applications usage duration' (indicating the time difference between the notifications created by smartphone applications categorized as Leisure), and 'Social media applications usage duration' (similarly, indicating the time difference between the notifications created by smartphone applications categorized as Social media.). The 'Sum of 213 other features' at the bottom aggregates the importance of all remaining features. The SHAP values here reflect each feature's average impact on predictions rather than indicating whether a feature's impact increases or decreases the likelihood of depression according to the model. It is important to note that these SHAP values indicate each feature's relative importance in this model's decision-making process and are not directly comparable to SHAP values from other models.

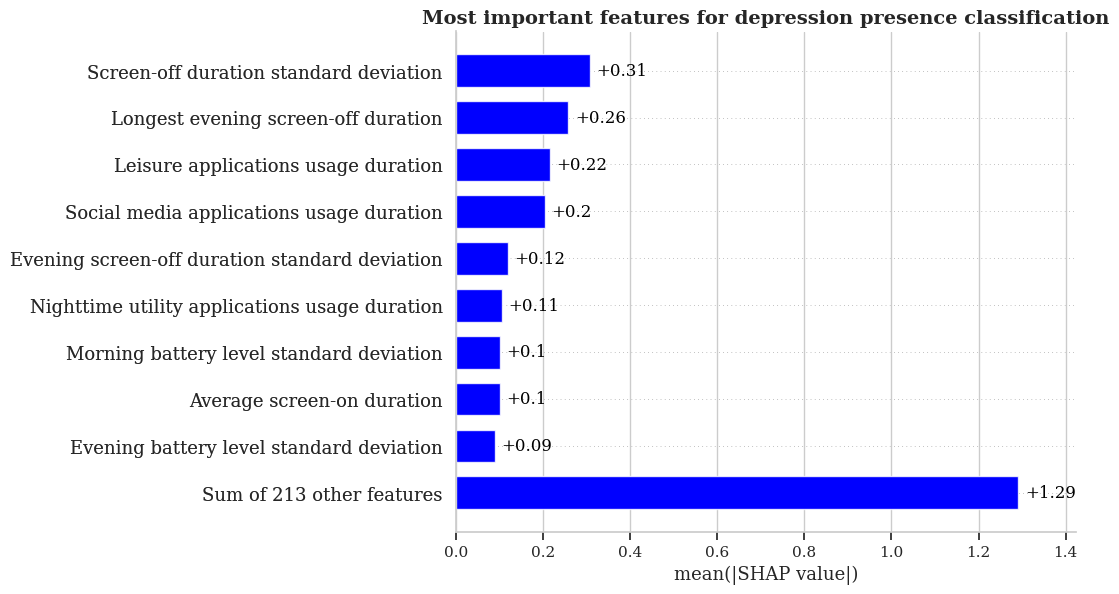

**Figure C2.** Key features in depression presence classification model with biweekly PHQ-9 score. This bar chart illustrates the most important features of a depression presence classification model (previous biweekly PHQ-9 score added as predictor), as evaluated using SHAP values. The 'Previous PHQ-9 Score' is the most impactful feature, followed by 'Standard deviation of screen-off duration' (indicating the fluctuation in smartphone screen-off durations), 'Longest screen-off duration' (indicating the longest period of smartphone screen turned off during the evening), and 'Average nighttime battery level' (indicating the average smartphone battery level during the night). The chart's base shows the collective impact of 202 other features. These SHAP values denote the average contribution of each feature to the model's predictions without specifying their direction of impact on depression likelihood. It is important to note that these SHAP values indicate each feature's relative importance in this model's decision-making process and are not directly comparable to SHAP values from other models.

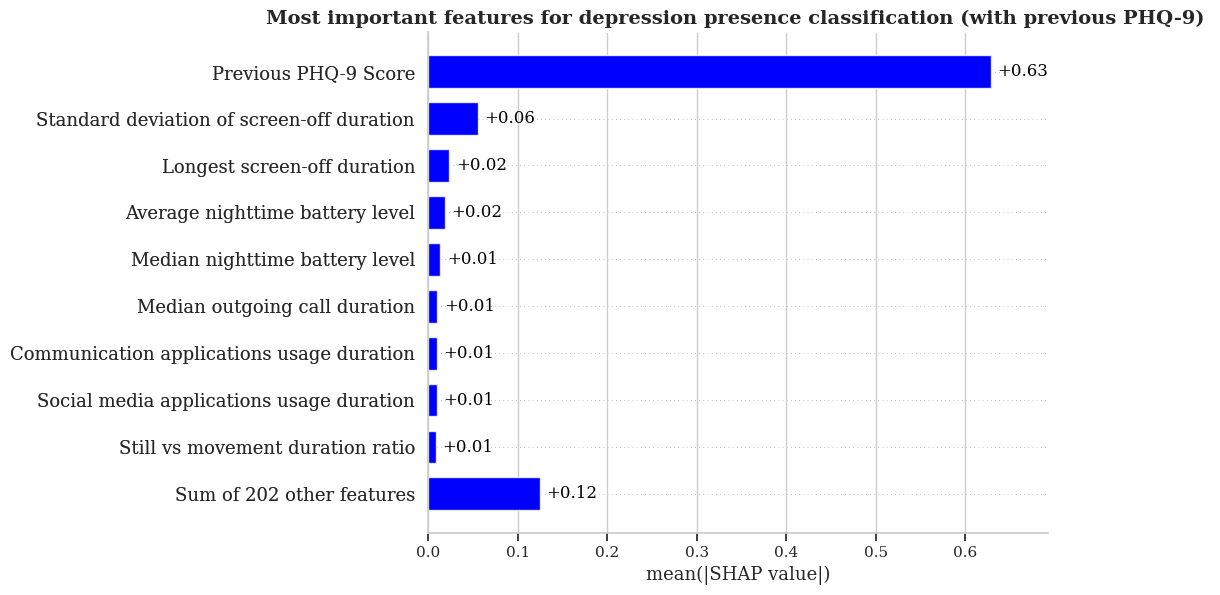

**Figure C3.** Key features in depression state transition classification with previous PHQ-9 score. This bar chart illustrates the most important features for a depression state transition classification model (previous PHQ-9 score added as predictor The 'Previous PHQ-9 score' emerges as the most important feature, followed by the 'Morning battery level standard deviation' (indicating smartphone battery level fluctuation during the morning), 'Acceleration magnitude count during the afternoon' (indicating the total movement of the subject, while carrying a smartphone, during the afternoon) and 'Maximum acceleration magnitude' (indicating the highest acceleration measured by the smartphone accelerometer). The features not explicitly shown in the table, indicated by the 'sum of 63 features', collectively have a less pronounced yet aggregated impact on the model's predictions. These SHAP values denote the average contribution of each feature to the model's predictions without specifying their direction of impact on depression likelihood. It is important to note that these SHAP values indicate each feature's relative importance in this model's decision-making process and are not directly comparable to SHAP values from other models.

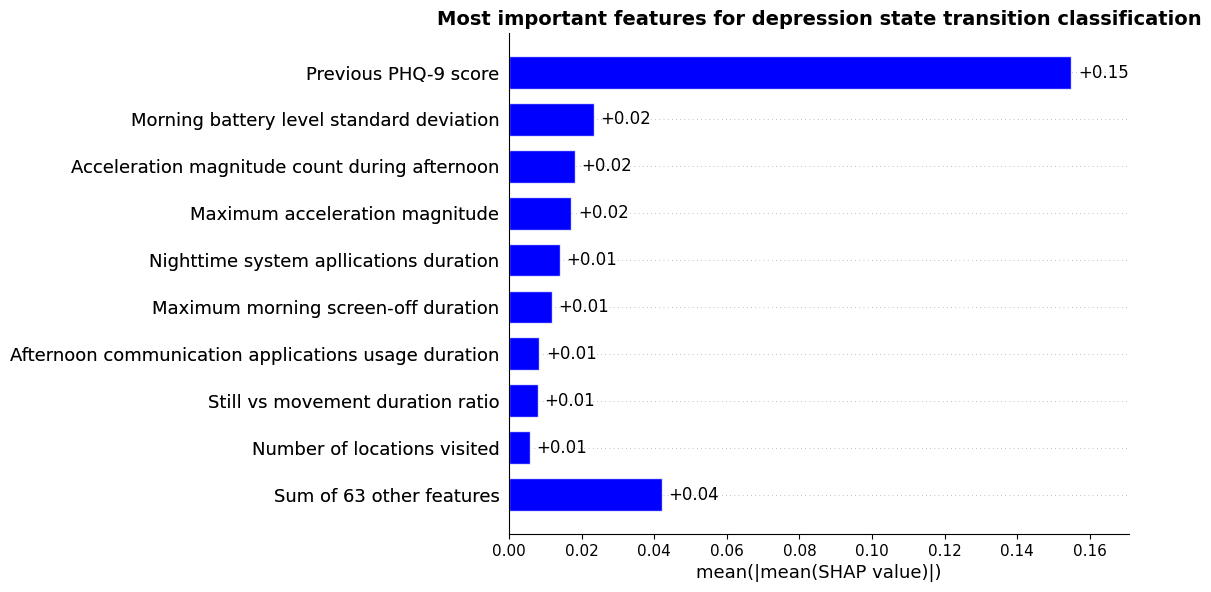
